# Region-based analysis of rare genomic variants in whole-genome sequencing datasets reveal two novel Alzheimer’s disease-associated genes: *DTNB* and *DLG2*

**DOI:** 10.1101/2021.06.09.21258576

**Authors:** Dmitry Prokopenko, Sanghun Lee, Julian Hecker, Kristina Mullin, Sarah Morgan, Yuriko Katsumata, Alzheimer’s Disease Neuroimaging Initiative (ADNI), Michael W. Weiner, David W. Fardo, Nan Laird, Lars Bertram, Winston Hide, Christoph Lange, Rudolph E. Tanzi

## Abstract

Alzheimer’s disease (AD) is a genetically complex disease for which roughly 30 genes have been identified via genome-wide association studies. We attempted to identify rare variants (minor allele frequency <0.01) associated with AD in a region-based, whole genome sequencing (WGS) association study (GSAS) of two independent AD family datasets (NIMH/NIA; 2247 individuals; 605 families). Employing a sliding window approach across the genome, we identified several regions that achieved p-values < 10^−6^, using the burden test or the SKAT statistic. The genomic region around the dystobrevin beta (*DTNB*) *gene* was identified with the burden test and replicated in case/control samples from the ADSP study (p_meta_ = 4.74×10^−8^). SKAT analysis revealed region-based association around the discs large homolog 2 (*DLG2*) gene and replicated in case/control samples from the ADSP study (p_meta_ =1×10^−6^). Here, in a region-based GSAS of AD we identified two novel AD genes, *DLG2* and *DTNB*, based on association with rare variants.

## Introduction

Alzheimer’s disease (AD) is a heterogeneous, genetically complex neurodegenerative disorder^1^. Genome-wide association studies (GWAS) have identified common variants in roughly 30 genes associated with AD^2,3^. GWAS heritability is estimated to be 24-33%^4,5^ - less than a half of the heritability calculated from twin studies^1^. Identification of rare variants associated with AD may help explain the missing heritability, and lead to new biological insights^6^. Several rare variant loci previously associated with AD^7^, including TREM2^8^, have been identified with whole-exome sequencing (WES) studies^9^.

Identification of association signals that are driven by rare variants remains cumbersome due to low power and relatively small sample sizes. Hence, aggregation methods, such as burden tests^10,11^ and variance component tests (SKAT)^12,13^, have been developed to jointly test regions of rare variants for association. Combining variant data increases the association signal and reduces the number of statistical tests. While burden tests are most powerful for signals with consistent effect directions, SKAT is more powerful for signals with different effect directions or when the fraction of causal variants is small. Previously, aggregated gene-based association analyses have been successful in identifying novel exome-wide significant associations with sporadic AD^14^.

Recently, a general framework for exact region-based association testing in family-based designs has been developed^15^. Using the proposed region-based testing framework, we performed a GSAS combining two AD family-based cohorts (605 families; 1509 affecteds; 738 unaffecteds) focusing on rare variants. For replication, we used case/control subjects from NIA ADSP, which included WGS data from a Non-Hispanic White (NHW) subcohort (983 cases; 686 controls), an African American (AA) subcohort (450 cases; 501 controls), and a Hispanic (HISP) subcohort (486 cases; 613 controls).

Using a p-value cutoff of 5×10^−6^, the burden test and SKAT identified several genomic regions showing association with AD risk. A region identified by the burden test in the *DTNB* gene (p=7×10^−8^) was replicated in the NHW samples. SKAT analysis revealed an association with variants encompassing a region around *DLG2* (p=4×10^−6^), which replicated in the NHW and the AA samples.

## Results

In a region-based whole-genome sequencing association study (GSAS) focusing on rare genomic variants, we combined two AD family-based cohorts, the NIMH Alzheimer’s disease genetics initiative study (NIMH) and the family component of the NIA ADSP sample. The combined sample consisted of 1509 affected and 738 unaffected siblings in families of predominantly European ancestry (Supplementary Table 1, Methods). 8,011,126 variants passed strict quality control and allele frequency (AF) filter of ≤1% (gnomAD^16^). We grouped rare variants into consecutive regions/windows of ten variants and performed a sliding-window rare variant WGS scan over the whole genome (801,124 windows). We employed a recently developed framework for exact regional-based analysis within FBAT^15^ to analyze these sets of rare variants using both the Burden test and SKAT. These tests are able to detect different configurations of disease regions - dense regions with the same effect directions (Burden test) or less dense signals with varying effect directions (SKAT).

Since we restricted our analysis to rare variants (i.e. AF <0.01) and given our modest sample size in the family-discovery cohort, we have used a relatively liberal p-value threshold p<5×10^−6^ to identify “suggestive associations” by burden test or SKAT. A stricter Bonferroni-corrected significance threshold would be p=6.24×10^−8^. Seven loci exhibited suggestive evidence for association with AD risk (Figure 1, Supplementary Figure 1, Table 1). For replication analysis, we selected the unrelated, multiethnic WGS AD subset from the NIA ADSP dataset (Methods). This dataset consists of three subpopulations: NHW (n=1669), AA (n=951), HISP (n=1099) (Sample sizes after quality control; Supplementary Table 1). A region located downstream to *DTNB*, with a Burden p-value of 7×10^−8^ in the discovery dataset, showed a burden p-value of 0.0324 in ADSP NHW (Table 1 and Supplementary Table 2). Another region, located in an intron of *DLG2* with a SKAT p-value of 4×10^−6^ in the discovery family-based dataset, showed replication with a significant SKAT p-value of 0.0143 in the ADSP NHW dataset and a SKAT p-value of 0.053 in the ADSP AA dataset (Table 1 and Supplementary Table 3).

**Figure 1:**
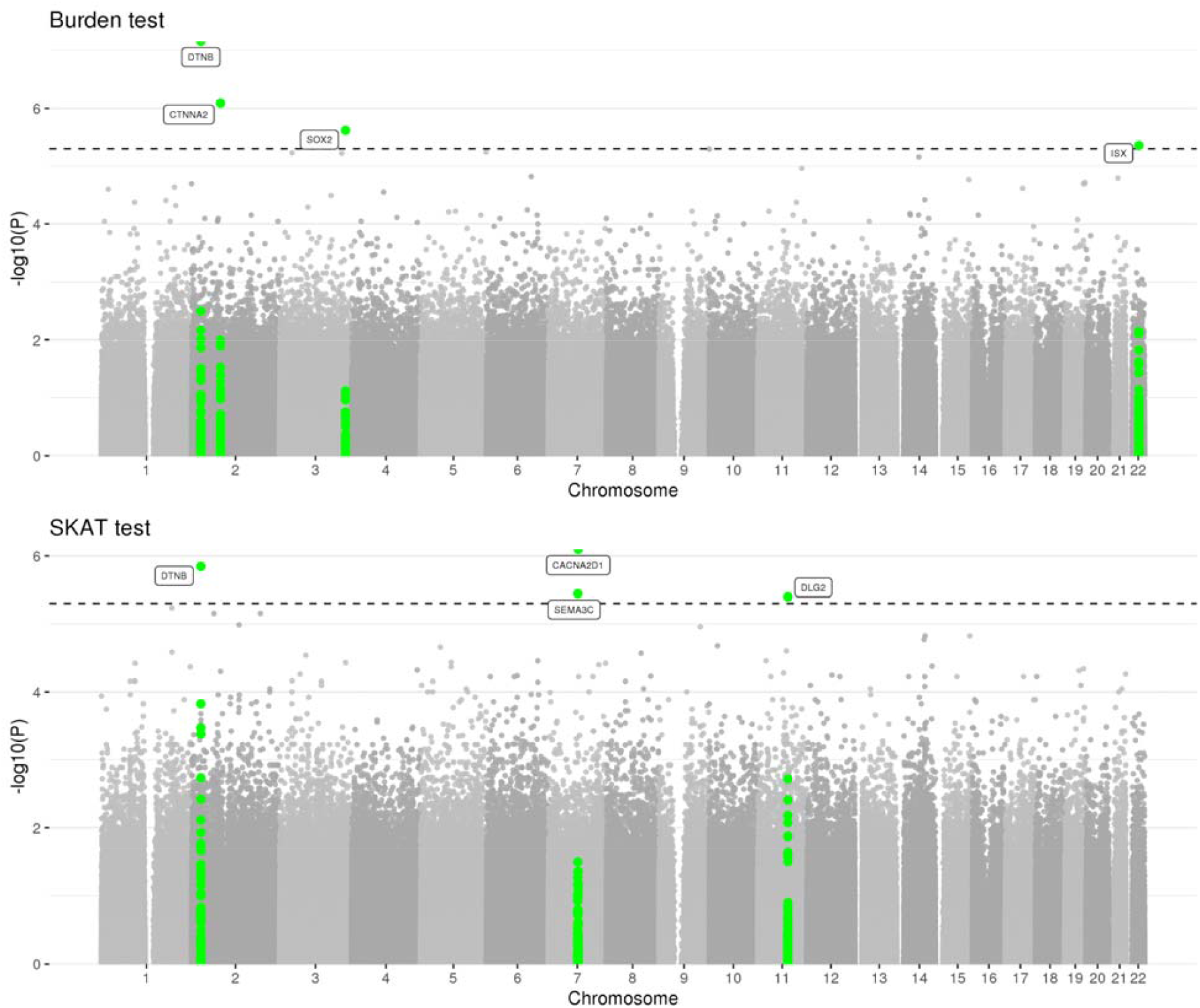
Manhattan plots of sets of rare variants in the whole genome scan of the family-based discovery dataset using the burden and SKAT test. Dashed line corresponds to suggestive threshold of 5×10^−6^.

**Table 1:** Top regions based on the burden or SKAT test with p<=5e-06 in the discovery family-based dataset using whole genome scan

| First SNV in the region<br>(chr:pos:ref:alt) | Last SNV in the region (chr:pos:ref:alt) | Nearest protein-coding gene | Discovery dataset (NIMH + NIA families) |  | Replication dataset NHW ADSP |  | Meta-analysis of family-based discovery and NHW ADSP replication datasets |  | Replication dataset AA ADSP |  | Replication dataset HISP ADSP |  |
| --- | --- | --- | --- | --- | --- | --- | --- | --- | --- | --- | --- | --- |
|  |  |  | P-value | nsim | P-value | Number of SNVs in the region | Fisher chi-squared test statistic | P-value | P-value | Number of SNVs in the region | P-value | Number of SNVs in the region |
| Burden test |  |  |  |  |  |  |  |  |  |  |  |  |
| 2:25703040:G:A | 2:25707419:T:G | DTNB | 7.00E-08 | 1.00E+08 | 0.032 | 5 | 39.808 | 4.74E-08 | 0.390 | 9 | 0.353 | 8 |
| 2:79854141:T:G | 2:79856252:C:T | CTNNA2 | 8.10E-07 | 1.00E+08 | 0.799 | 6 | 28.500 | 9.88E-06 | 0.774 | 8 | 0.785 | 7 |
| 3:181942653:A:G | 3:181946475:G:A | SOX2 | 2.40E-06 | 1.00E+07 | 0.317 | 6 | 28.177 | 1.15E-05 | 0.619 | 9 | 0.163 | 8 |
| 22:35048628:G:C | 22:35053269:C:T | ISX | 4.40E-06 | 1.00E+07 | 0.766 | 8 | 25.201 | 4.58E-05 | 0.827 | 9 | 0.014 | 10 |
| Variance component (SKAT) test |  |  |  |  |  |  |  |  |  |  |  |  |
| 11:83498255:A:G | 11:83500398:T:G | DLG2 | 4E-06 | 1.00E+07 | 0.014 | 5 | 33.352 | 1E-06 | 0.053 | 8 | 0.893 | 9 |
| 2:25703040:G:A | 2:25707419:T:G | DTNB | 1.4E-06 | 1.00E+08 | 0.054 | 5 | 32.737 | 1.4E-06 | 0.409 | 9 | 0.455 | 8 |
| 7:82268137:T:C | 7:82271095:A:T | CACNA2D1 | 8E-07 | 1.00E+07 | 0.591 | 5 | 29.131 | 7.4E-06 | 0.278 | 8 | 0.090 | 8 |
| 7:81141368:T:G | 7:81143780:C:T | SEMA3C | 3.6E-06 | 1.00E+07 | 0.251 | 5 | 27.832 | 1.4E-05 | 0.046 | 9 | 0.694 | 10 |

SNV - single nucleotide variant, NHW - non Hispanic white, AA - African-American, HISP - Hispanic

Both *DLG2* and *DTNB* are highly expressed in the brain based on RNA-data from three different sources: Internally generated Human Protein Atlas (HPA) RNA-seq data, RNA-seq data from the Genotype-Tissue Expression (GTEx) project, and CAGE data from the FANTOM5 project, as well as the consensus dataset for each gene derived from the Human Protein Atlas^17^ (Supplementary figures 2 and 3). In the Alzheimer’s Disease Dataset analysis^18^ (GSE48350) from the GEO database^19^ expression of *DLG2* and *DTNB* is significantly decreased in AD compared to control subjects (Supplementary table 4).

Network analysis revealed a network of 33 proteins interacting with DLG2 and DTNB that were enriched for neuronal synaptic functions (Supplementary Figure 4). Functional enrichment of the subnetwork of proteins directly interacting with DLG2 and DTNB revealed 694 enriched GO process/ pathway terms (Supplementary table 5). The most enriched part of the network was for proteins interacting with DLG2 that are connected to neurexins and neuroligins, as well as trafficking of AMPA receptors. DLG2 also interacted with 4 proteins (NOS1, ERBB4, DLGAP2, NRXN3) previously associated with AD risk^20^, and 4 proteins (GRIN1, GRIN2A, GRIN2B, GAPDH) associated with AD in the KEGG Alzheimer’s pathway. DLG2 and DTNB also share protein-protein or co-expression interactions through KIF1B, MLC1, and SH3D19.

## Discussion

Here, we describe a comprehensive region-based analysis of Alzheimer’s disease using WGS datasets. We specifically searched for novel AD association signals driven by regions of rare variants in a large family-based cohort. To account for different disease region specifications, we employed both the burden test and SKAT. This yielded seven regions of suggestive evidence (p<5×10^−6^) for association with AD risk in the family datasets. These results were followed up with replication analysis in independent case-control samples of different ethnicities. Two loci, *DTNB* and *DLG2*, showed consistent evidence of replication in at least one of the subpopulations (i.e. NHW). The *DLG2* region was also confirmed in the African American sample.

*DLG2* encodes a member of the membrane-associated guanylate kinase family, also known as post-synaptic density protein, PSD-93. Down-regulation of synaptic scaffolding proteins, including DLG2, has been described as an early event in AD^21^. DLG2 has been proposed as a potential target for AD based on an integrated metabolomics-genetics-imaging systems approach in Agora (URLs); agonism of DLG2 is predicted to reduce disease progression. An expression dataset of AD in the GEO database revealed reduced expression of *DLG2* in AD versus controls. A common variant in *DLG2*, rs683250, was previously associated with increases of shape asymmetry in controls as compared demented populations^22^. This same variant is in linkage disequilibrium (LD, D’=1) with all rare variants of the *DLG2* region found to be associated with AD here. *DLG2* variant, rs286043 (AF=0.03), which exhibited suggestive evidence for association with AD risk in IGAP (p=5e-06), is in LD with 4 out of 10 variants from our *DLG2* AD-associated region, suggesting possible allelic heterogeneity. *DLG2* has previously been associated with schizophrenia^23^ and autism^24,25^. Along these lines, *DLG2* deficiency in mice has been reported to lead to reduced sociability and increased repetitive behavior along with aberrant synaptic transmission in the dorsal striatum^26^.

β-Dystrobrevin (*DTNB*) is associated with neurons in the cortex, hippocampus, and cerebellum, and has also been reported to be enriched in the post-synaptic density^27^. Kinesin superfamily motor proteins (KIF) are responsible for anterograde protein transport within the axon of various cellular cargoes, including synaptic and structural proteins^28^. Dysregulated KIF expression has also been associated with early AD pathology^29^, and β-Dystrobrevin interacts directly with kinesin heavy chain in the brain^30^. Dystrobrevin-binding protein 1, also known as dysbindin, has been reported to be associated with schizophrenia^31,32^. Thus, both novel AD gene candidates identified in this study have been associated with post-synaptic function and schizophrenia.

Our approach utilized two region-based tests (burden and SKAT) in a family-based design, in which the joint distribution of rare variants is not estimated, but rather obtained by the haplotype algorithm for FBAT, which is robust against population structure and admixture, and allows for construction of exact or simulation-based p-values. Previously, we performed region-based rare variant testing, but with different region definitions, and using only burden tests with empirical estimation of the variant correlations and asymptotic p-values^20^. While this is the largest combined family-based AD-specific WGS dataset available, larger datasets will be needed to confirm our findings in future studies. We also note that by utilizing a window size of 10 consecutive variants, we could have missed sparsely distributed signals. Since the number of possible haplotypes increases exponentially with the number of variants tested, larger window sizes were computationally infeasible.

In summary, we identified two novel loci associated with AD, based on association with rare variants in *DLG2* and *DTNB* in a family-based AD WGS sample using methods that are robust to population structure. We further showed replication in an independent multi-ethnic AD WGS dataset with unrelated cases and controls. These findings demonstrate the usefulness of WGS in capturing non-exonic, rare variant signals. Both novel AD-associated genes identified here encode post-synaptic density proteins and have been implicated for roles in schizophrenia.

## Data Availability

Data available upon request. Summary statistics will be made available after peer-review.

## Acknowledgements

This work was supported by Cure Alzheimer’s Fund and NIH R56AG057191 (D.W.F. and Y.K.). The computations in this paper were run in part on the Odyssey cluster supported by the FAS Division of Science, Research Computing Group at Harvard University with support from John Morrissey and in part on compute provided by Dell HPC Research Computing Solutions with support by Glen Otero. The funding body has no role in the design of the study and collection, analysis, and interpretation of data and in writing the manuscript. Please refer to the Supplementary Note for full acknowledgements.

## Author Contributions

D.P., C.L., R.E.T. contributed to the study concept and design. D.P., S.L., K.M., S.M., Y.K., D.W.F., L.B., W.H., C.L., R.E.T. contributed to data analysis and/or interpretation. J.H., N.L., C.L. contributed to statistical support. D.P., S.L., W.H., C.L. and R.E.T. wrote the original draft of the paper, and all authors critically reviewed the manuscript.

## Methods

### Study populations

#### Discovery family-based dataset

Our discovery dataset consisted of two WGS family-based cohorts: the National Institute of Mental Health (NIMH) family AD cohort^33^ and families from the National Institute of Aging Alzheimer’s Disease Sequencing Project^34^ (NIA ADSP). Whole-genome sequencing and variant calling in NIMH are described elsewhere^35^. Variant calls for the families from the NIA ADSP cohort were obtained from the National Institute on Aging Genetics of Alzheimer’s Disease Data Storage Site (NIAGADS) under accession number: NG00067. Both cohorts consisted of multiplex AD families with affected and unaffected siblings (Supplementary table 1). A subject was considered to be affected if he/she was included in one of the following categories: “Definite AD”, “Probable AD” or “Possible AD”. Unaffected subjects had either no dementia, suspected dementia (46 subjects), or non-AD dementia (10 subjects). It is important to note that NIA ADSP families by design did not include individuals with two APOE-ε4 alleles. After standard quality control, both cohorts were merged together.

#### NIA ADSP case-control dataset

WGS variant calls for the NIA ADSP replication case-control dataset were obtained from the NIAGADS under accession number: NG00067 and consisted of the ADSP Discovery-Extension Case-Control WGS dataset^34^ and the ADNI Case-Control WGS dataset. Samples were remapped to GRCh38 and jointly called with the families from the NIA ADSP cohort. Full details can be found on NIAGADS (https://dss.niagads.org/datasets/ng00067/) and elsewhere^36^. Briefly, a subject was considered affected, if he/she met the NINCDS-ADRDA criteria for possible, probable, or definite AD, had documented age at onset or age at death (for pathologically verified cases), and APOE genotyping. All controls were 60 or more years old and were free of dementia.

### Quality control

Briefly, we have excluded individuals based on genotyping rate, inbreeding coefficient, and family mismatches using identity by descent (IBD) sharing coefficients. After sample-based quality control, we have combined two WGS family-based cohorts NIMH (1,393 individuals in 446 families) and 854 individuals (families from NIA ADSP; 159 families). In the merged dataset we excluded multiallelic variants, monomorphic variants, singletons (i.e. variants with only one alternative allele across the dataset and variants seen only in one family), indels, and variants which had one missing allele among 2 alleles in an individual. The remaining variants were filtered based on Mendel errors, genotyping rate (95%), Hardy-Weinberg equilibrium (p<1e-08), calling quality in TOPMed, which is a large WGS database with >100,000 individuals sequenced jointly, and allele frequency in gnomAD (AF<= 1% in either whole gnomAD or nonFinnish European sample).

### WGS regional-based analysis

We have performed a whole genome scan for our combined family-based AD dataset using a newly developed exact framework in FBAT for region-based association testing^15^. We grouped rare variants in consecutive sets of ten. For each set of rare variants, we considered the burden test and the SKAT test using Affection Status minus offset as phenotype. We selected an offset of 0.15 which approximately corresponds to the population prevalence of AD. We have used FBAT^37^, R^38^, snakemake^39^ and bash commands to implement and run the described analyses.

### Replication

Replication significance level was set to 0.0143 (0.1 divided by 7 independent loci). Regions/windows with P<=0.05 were also reported as replicated. We have used the SKAT package to perform Burden and SKAT-O tests on the same sets of rare variants in the case-control replication cohorts. As covariates, we used sequencing center, age, sex, and principal components (to account for population structure). Principal components were calculated based on rare variants using the Jaccard index^40^. We have also performed meta-analysis among datasets with similar ethnical background using the Fisher’s combined probability test.

### RNA-Seq and microarray analysis

We explored DLG2 and DTNB genes’ expression based on the Human Protein Atlas (HPA) RNA-seq data (https://www.proteinatlas.org) and tested for differential expression of synaptic and immune related genes including *DLG2* and *DTNB* genes between normal controls (N=173, aged 20-99 yrs) and AD cases (N=80) in the brain regions including hippocampus, entorhinal cortex, superior frontal cortex, and post-central gyrus using microarray dataset GSE48350, which is available from the Gene Expression Omnibus Web site (http://www.ncbi.nlm.nih.gov/geo/). Differential expression was tested using the “GEO2R” tool.

### Network construction

We used Cytoscape 3.8.0 and the StringDB protein-protein interaction resource^41^ using only identified protein-protein interactions. Using a background that agglomerates protein-protein interaction datasets, we seeded the network with DLG2 and DTNB and identified direct associations between proteins and DLG2 and DTNB in a global network (supplementary table 5). Results were combined using the Genemania server (Utilizing significantly co-expressed genes across several experimental datasets)^42^ to further capture functional relationships and to build a combined protein-protein/gene co-expression network.

### Functional enrichment

Functional enrichment within the network was performed using the remote StringDB server linked to Cytoscape “String App Enrichment function”^43^, producing enrichments using the hypergeometric test, with *P*-values corrected for multiple testing using the method of Benjamini and Hochberg in known molecular pathways and GO terms as described in Frenceschini *et al*.^44^

## URLs

FBAT, https://sites.google.com/view/fbatwebpage; gnomAD, https://gnomad.broadinstitute.org/; Agora AMP-AD, https://agora.ampadportal.org/genes; TOPMED, https://www.nhlbiwgs.org/; Human Protein Atlas, https://www.proteinatlas.org/; GEO database, https://www.ncbi.nlm.nih.gov/geo/; NIAGADS, https://www.niagads.org/; StringDB, https://string-db.org/.

